# Characteristic and Sex Differences in Auditory Function and Cochlear Pathophysiology in a Noise-exposed Cohort: A Cross-sectional Study

**DOI:** 10.1101/2020.09.27.20202481

**Authors:** Li Bei, Wang Qixuan, Yang Lu, Li Yun, Huang Zhiwu, Wu Hao

## Abstract

**Background:** To determine the characteristics and sex differences of auditory perception and cochlear function in individuals with long-term occupational noise exposure.

**Methods:** Young workers with long-term occupational noise exposure from a shipyard were recruited in the current study as the hidden hearing loss (HHL) risk group. Age-matched office workers in the same shipyard who had no occupational noise-exposure history were enrolled in the control group. The auditory processing ability of speech-in-noise (SIN) score and gap detection threshold (GDT) were further examined by sex. The cochlear function of action potential (AP) and summating potential (SP)/AP values were tested and compared by sex and side. The correlation between the SIN score and cochlear function was studied by sex. The correlation between either auditory processing ability or cochlear function and occupational-noise working length (OWL) was also analysed in the HHL risk group.

**Results:** Significantly decreased SIN scores and a higher GDT of the 4 kHz gap marker were only found in men in the HHL risk group. Although the hearing thresholds of the women in the HHL risk group were slightly but significantly worse than those of the women in the control group, no significant defects in auditory processing or temporal resolution were found between the two groups. Significantly decreased cochlear function and increased SP/AP values in the left ear were only found in men in the HHL risk group. Neither the AP amplitude nor the AP latency differed significantly between the two groups by sex. A correlation study indicated that only the correlation between the SIN score and the AP amplitude of the right ear in men was significant. No significant difference was found between the SIN score and cochlear function in women. The AP latency of the right ear was only significantly correlated with OWL in men.

**Conclusion:** In long-term occupational working exposure individuals with normal hearing, defects in auditory processing, temporal resolution and cochlear function showed sex differences, none of which were significant in women. In men, a weak correlation between the SIN score and the AP amplitude of the right ear was found. There was only a weak correlation between OWL and the AP latency of the right ear in men. Our findings indicate men are more vulnerable to occupational noise than women. Considering the noise-exposure dose differences between the control and HHL risk groups, our measures are insensitive to cochlear synaptopathy in humans.

## Background

Exposure to hazardous noise is one of the most common occupational risks worldwide. It has been suggested that 12% or more of the global population is at risk for hearing loss from noise, equating to well over 600 million people [1]. Noise exposure could lead to hearing impairment, irritability, disturbed sleep, daytime sleepiness, worse patient outcomes and staff performance in hospitals, and an increased incidence of hypertension and cardiovascular disease [2]. Noise-induced hearing loss is one of the most common diseases in people exposed to occupational noise. Initial studies discovered that there was a characteristic “notch” in hearing ability at 4000 Hz and showed that the frequency, intensity, and duration of the exposure were also key factors that influenced the degree of hearing loss sustained [3, 4]. As a result, monitoring the hearing threshold, especially between 3000 Hz and 6000 Hz, has become routine among individuals exposed to occupational noise.

The consequences of noise-induced damage is still an ongoing debate. Animal studies have shown that moderate noise could induce a temporal threshold shift (TTS) and cause damage to the synapse between the inner hair cells (IHC) and type I afferent auditory nerve fibres [5-7]. This kind of pathology is termed cochlear synaptopathy (CS). This form of hearing loss has also been referred to as “hidden hearing loss” (HHL) to reflect that the dysfunction is not revealed by standard tests of auditory thresholds.

Along with synapses and afferent auditory nerve fibre injury, other coding deficiencies, including loudness and temporal resolution, are also observed. For instance, Shi et al. [8] found that the click-evoked compound action potential (CAP) amplitude was reduced after noise exposure in guinea pigs. Song et al. [9] showed that the click-evoked CAP amplitude decreased with prolonged peak latency. Other studies also reported a significant reduction in the auditory brainstem response (ABR) I wave amplitude in CS rodents [5, 6, 10].

The discovery of HHL has challenged the long-held belief that noise exposures that do not result in audiometric threshold shifts are safe. Many human subjects show normal auditory thresholds but have significant perceptual difficulties, including understanding speech in noisy backgrounds. Animal studies suggest that CS could also play an important role in these populations. However, CS can only be effectively studied in animal models with a combination of invasive physiological and histological tests, which is not possible in human subjects. Therefore, specific, sensitive, and reliable noninvasive diagnostic tests are essential. Some electrophysiological values, such as the ratio of summating the potential (SP) relative to the action potential (AP) [11], the ABR Wave I amplitude [12, 13], the ABR wave V latency [14] and the frequency-following response (FFR) [15], have been reported as indicators of CS. Some of the electrophysiological indicators showed that CS and auditory nerve fibre degeneration also occurred in exposed populations. For example, Stamper and Johnson[12, 16] found that the ABR wave I amplitude decreased as a function of noise exposure amount only in women. Liberman et al. [11] reported that the SP/AP value increased in the HHL high-risk group. However, Prendergast found no relationship between noise exposure and the amplitude of the ABR [17]. It is important to note that controversy still exists regarding whether CS occurs in humans.

If noise-induced CS does exist in humans, it is most likely be measurable in humans with extreme noise exposure. Although the long-term, continuous exposure at work is somewhat different from the one-off acoustic trauma in animal studies, it could be one of the extreme cases of noise exposure in humans, and CS could possibly be seen in individuals with long-term occupational noise exposure. To explore the effect of long-term occupational noise exposure on the human auditory system, young workers with long-term occupational noise exposure from the Production Department of a shipyard were recruited as the HHL risk group in this study. Age-matched workers from the logistics, purchasing and accounting department in the same shipyard with no occupational noise exposure were enrolled as the control group. To exclude any effect of sex, individuals enrolled in each group were further divided by sex. Comparisons were conducted between the male and female groups. Both the SIN score and gap detection threshold (GDT) were investigated. The AP and SP/AP values were recorded and compared. The correlation of the SIN score with cochlear function was analysed by sex. Finally, the correlation of OWL between either auditory processing ability or cochlear function was studied in the HHL risk group. The purpose of this study was as follows: (1) to explore whether auditory processing disorder and cochlear dysfunction appear in individuals exposed to long-term noise, (2) to study the correlation of speech-in-noise (SIN) scores and cochlear function by sex and (3) to study the effect of occupational-noise working length (OWL) on auditory processing disorder and cochlear dysfunction by sex.

## Methods

### Study design and participants

This was a cross-sectional observational study performed from August to October 2019 and it was approved by the Translational Medicine Ethics Review Committee of Shanghai Ninth People’s Hospital Affiliated to Shanghai Jiao Tong University, School of Medicine. All participants signed an informed consent form before participating.

### Participant recruitment

The male and female individuals in the HHL risk group and the control group were recruited from the same factory of a shipyard. The annual hearing test reports of all of the workers were reviewed before recruitment. Only individuals with normal hearing reports were recruited. Individuals from the production department were screened for inclusion in the HHL risk group. Individuals from the logistics, purchasing and accounting department with little or no occupational noise were enrolled in the control group.

Individuals were instructed to fill out a questionnaire, and gender, age, handedness, auditory disease and ototoxicity history, working department, working hours per week and occupational-noise working duration were collected. OWL was defined as the months of working in the occupational-noise environment and was calculated as years by dividing by 12. An otoscope examination was performed to check the external auditory canal and tympanic membrane of both ears. Hearing thresholds of both ears were reevaluated in all participants. Pure-tone thresholds of 250, 500, 1000, 2000, 3000, 4000, 6000, and 8000 Hz were tested.

Enrolment criteria in the HHL risk group included (1) age between 20 and 40 years old, (2) no ear disease or ototoxicity history, (3) hearing threshold of 250, 500, 1000, 2000, 3000, 4000, 6000, and 8000 Hz at less than or equal to 25 dB HL, and (4) more than or equal to 3 working years in the production department. The enrolment criteria in the control group were the same as those in the HHL risk group except that they had no occupational noise exposure history or shooting habits.

## Auditory processing ability

### Speech-in-noise test

The original speech material was the Mandarin version of the Hearing in Noise Test (MHINT). MHINT contains 14 lists that contain 20 sentences each. Each sentence contains 10 keywords. The scores are expressed as percentages of the keywords that were heard correctly. The MHINT sentences were recorded with a male speaker. The stimuli were presented at 65 dB SPL (A) and were delivered bilaterally through Sennheiser HD580 headphones. In sentence recognition above noise, speech-shaped noise was presented at 65 dB SPL (A) bilaterally. The noise began 500 ms before the sentence and continued for 500 ms after the sentence had finished.

All participants had no experience with any speech tests before this study. Before the formal test, participants practiced 3 to 5 times with each sentence and were provided feedback to become familiar with the stimuli. In the formal test, they were instructed to repeat the sentences as accurately as possible. Each sentence was played only once, and no feedback was provided during the formal tests.

### Gap detection threshold test

The GDT test was measured in a three-interval forced-choice procedure. For the gap marker, white noise was low-pass filtered at cut-off frequencies of 1, 2, and 4 kHz via a 3000th-order finite impulse response filter with an approximately −116 dB/octave filter slope. In brief, a three-interval forced-choice program was run on MATLAB software (version 7.0). Three buttons were presented on a monitor to the participant who was asked to indicate which one of the three stimuli were different (i.e., which of the three stimuli was inserted with a gap). Details of the GDT test may be found in Li et al. 2017 [18].

## Cochlear function

### Electrocochleography recording

Electrocochleography recordings were collected using a commercial device (Intelligent Hearing Systems, US) with Smart EP software. The electrode impedance values were all less than 5 kΩ, and the interelectrode impedance was within 1 kΩ. A silver electrode with cotton and electrode gel was applied as the reference electrode in the ear canal. An electromagnetically shielded insert earphone (ER-3) was applied to deliver click stimulation to the test ear at 90 dB nHL in alternating polarity at a rate of 7.1/sec. The recorded potentials were amplified by a factor of 50,000 and filtered with 10 Hz (high-pass) and 3000 Hz (low-pass) filters. Averaged responses over 512 sweeps were acquired. The AP waves of both ears were marked. The amplitude and latency of AP in both ears were recorded. The SP/AP value for either ear calculated automatically by the machine was recorded and analysed.

## Correlation analyses

The correlation of the SIN score and the indicators of cochlear function was studied by group and sex. The correlation between either auditory processing ability or cochlear function and OWL was calculated for the HHL risk group.

## Statistics

Data were analysed before applying statistics and graphing. Data values beyond the mean±3 SD were considered outliers and were excluded. Individuals in the different groups were further divided by sex. Comparisons were performed in the male control, female control, male HHL risk and female HHL risk groups. To reduce the Type I error in statistics in the four groups, analysis of variance (ANOVA) or the Kruskal–Wallis H nonparametric test was applied. Data distribution was checked before statistical analysis in the two groups. If the data complied with the normal distribution, ANOVA was performed. Otherwise, the Kruskal–Wallis H test was applied. The Bonferroni method was applied for post hoc tests in ANOVA, and only the results of the same sex in different groups are displayed. All statistical analyses were performed using SPSS (IBM SPSS Statistics version 20). p values less than 0.05 indicated significance.

## Results

### Demographic characteristics and the audiometry of the participants

In total, 142 individuals were recruited and assessed for eligibility. Among these individuals, 120 met the criteria and were included in the study. There were 60 participants (27 women) in the HHL risk group and 60 participants (24 women) in the control group. OWL in the HHL risk group ranged from 3 to 15.08 years, mean (SD) 7.4±2.78 year. The mean (SD) ages of the *male* participants were 27.6 (4.66) years for the control group and 27.9 (3.37) years for the HHL risk group. The mean (SD) ages of the *female* participants were 28.0 (3.94) years for the control group and 28.5 (4.08) years for the HHL risk group. ANOVA showed that the age differences among the four groups was not significant (F = 0.231, p = 0.874).

Figure 1 shows the audiograms of the participants in the control group and the HHL risk group. The mean (SD) hearing threshold of the left ear for *men* in the control group was 9.7 (2.95) dB HL, and that in the HHL risk group was 10.0 (3.63) dB HL. The hearing threshold of the right ear for *men* in the control group was 10.1 (2.60) dB HL, and that in the HHL risk group was 11.3 (3.07) dB HL. The hearing threshold of the left ear for *women* in the control group was 6.4 (3.55) dB HL, and that in the HHL risk group was 10.1 (2.81) dB HL. The hearing threshold of the right ear for *women* in the control group was 8.1 (3.63) dB HL, and that in the HHL risk group was 11.7 (3.20) dB HL. ANOVA showed that the hearing differed significantly between the left and right side in all four groups (both p < 0.001). Post hoc tests showed that the hearing threshold for the *men* in the two groups was not significant (p*left* = 1.000, p*right* = 0.725). The hearing threshold for the *women* in the two groups was significant (p*left* = 0.001, p*right* < 0.001).

**FIGURE 1.**
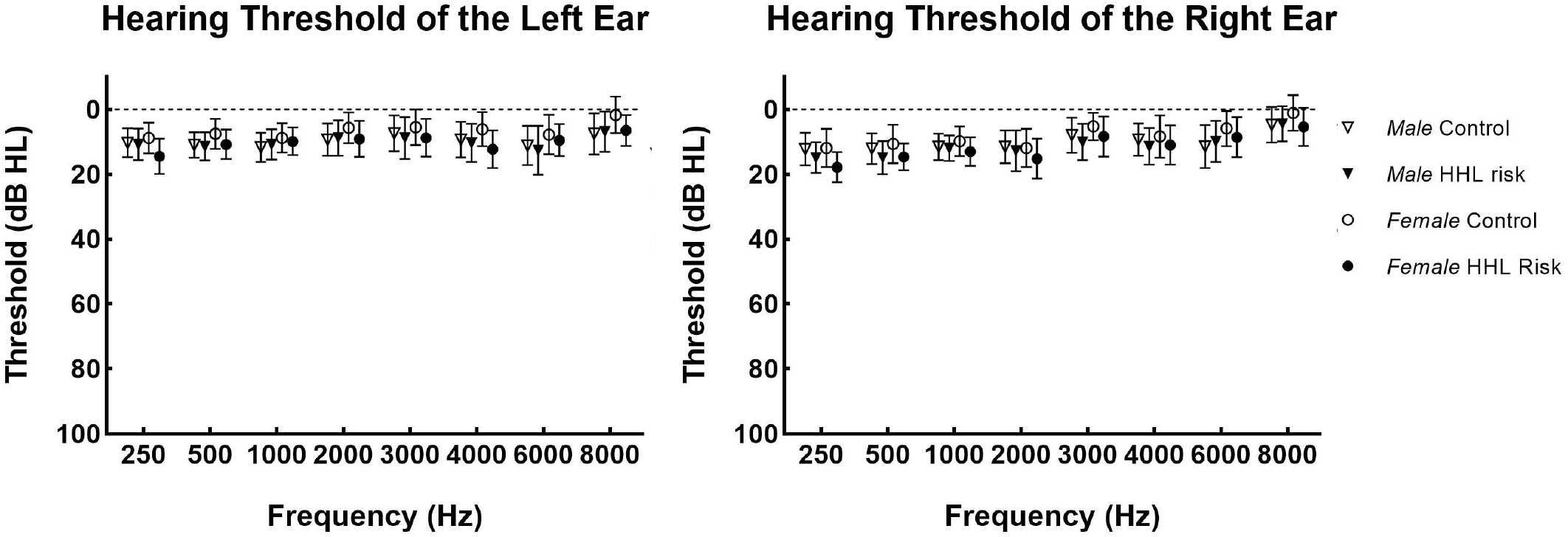
Pure tone audiometric thresholds. Hearing thresholds (averaged across male and female listeners) are shown by ear, with mean and standard errors in different panels. Significance was found between the control and HHL risk groups in women (data not shown).

### Auditory function feature of the two groups

#### Speech-in-noise

SIN scores for individuals in either group are shown in Figure 2. In the noise condition (SNR = 0), both the control group and the HHL risk group achieved high scores. The mean (SD) scores of the *male* participants were 96.13 (2.322) for the control group and 93.20 (3.883) for the HHL risk group. The mean (SD) scores of the *female* participants were 95.10 (1.994) for the control group and 92.26 (4.404) for the HHL risk group. A nonparametric test showed that the differences among the four groups were significant (p < 0.001). Post hoc tests showed that a significant difference was shown between the *men* in the control group and the HHL risk group (p = 0.005). The differences between the *women* in the control group and those in the HHL risk group were not significant (p = 0.126) (Fig. 2).

**FIGURE 2.**
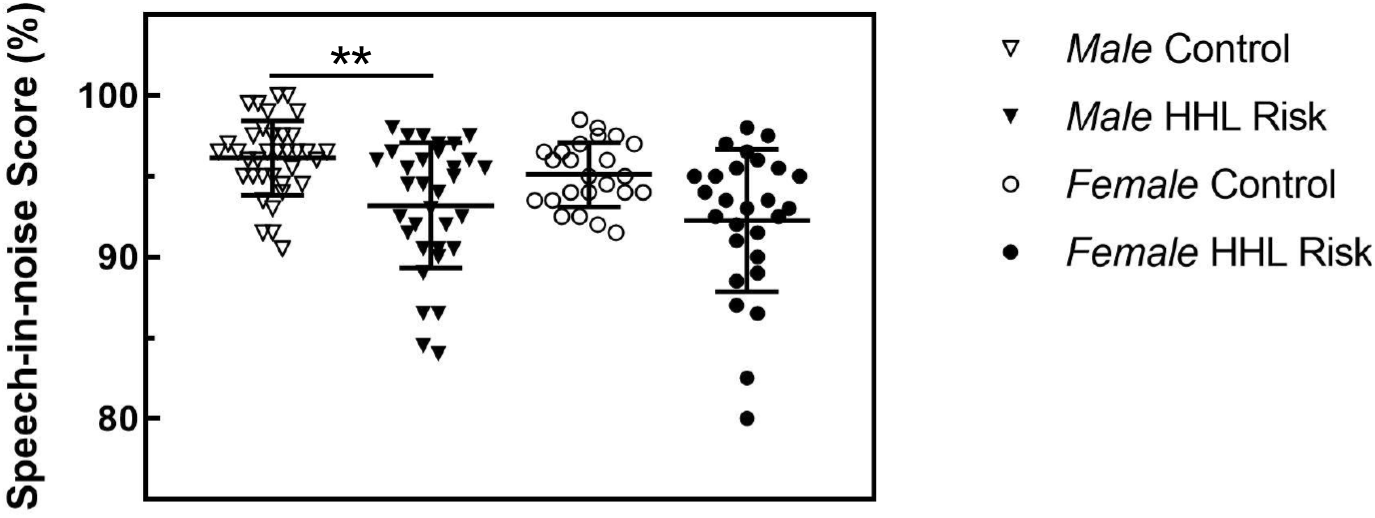
SINs for the control and the HHL risk group by sex. Mean and standard errors are calculated by sex and group. Significant differences were found between the control and HHL risk groups in men. The p values less than 0.01 are indicated **.

### Gap detection threshold

The GDTs in the groups varied in terms of gap marker cut-off frequency, as shown in Figure 3. Generally, the GDTs of the control group were lower than those of the HHL risk group with the same gap marker. The mean (SD) threshold of *men* in the control group was 2.79 (0.629), 4.04 (0.903) and 6.12 (1.506) for the 4 kHz, 2 kHz and 1 kHz markers, respectively. Those in the HHL risk group were 3.57 (1.273), 4.22 (1.073) and 6.77 (1.942). The mean (SD) thresholds of the *women* in the control group were 2.53 (0.337), 3.80 (0.828) and 5.77 (1.267) for the 4 kHz, 2 kHz and 1 kHz markers, respectively. Those in the HHL risk group were 3.15 (1.243), 3.93 (1.174) and 6.24 (1.245). Data derived from subjects with the same gap marker frequencies were analysed. A nonparametric test was applied in the analysis of the 4 kHz gap marker, and ANOVA was performed for those of the 2 kHz and 1 kHz gap markers. A significant difference was observed for the 4 kHz gap marker in *men* (p = 0.042). A nonparametric test showed that the difference between the control and HHL risk groups at the 4 kHz gap marker in *women* was not significant (p = 0.255). No significant difference was evident at the *2 kHz (F2 kHz* = 0.874, p = 0.457) and 1 *kHz gap markers (F1 kHz* = 1.988, p = 0.120) (Fig. 3).

**FIGURE 3.**
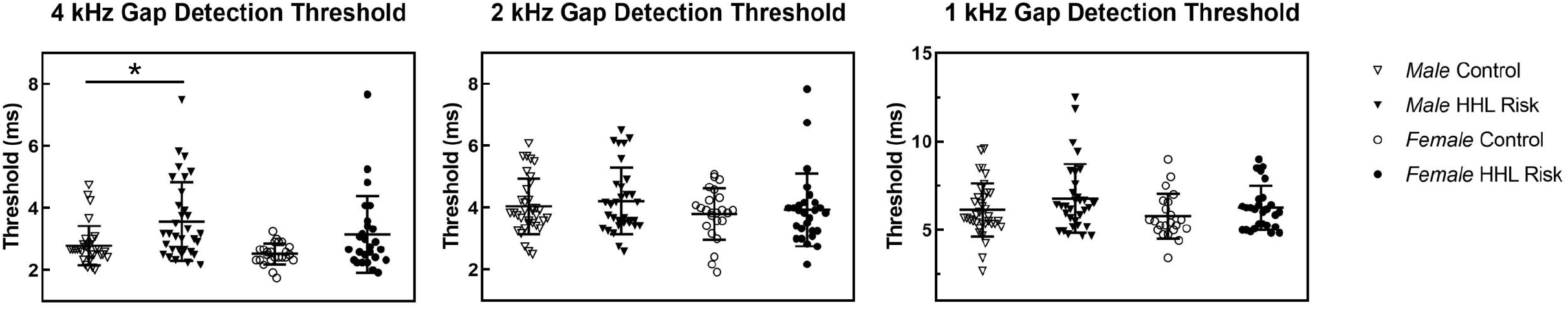
The GDTs for the control and HHL risk groups by sex. The GDTs at different gap markers are shown in different panels. GDTs further separated by sex were compared. Significance was only found in men with a 4 kHz marker. The p values less than 0.05 are indicated *.

### Cochlear function feature of the two groups

#### Electrocochleography

The values of the AP amplitude of both ears are plotted in Figure 4. The details of the *left* ear are plotted on the left, and those of the *right* ear are plotted on the right. The mean (SD) values of the *left* AP amplitude in *men* were 0.86 (0.434) for the control group and 0.91 (0.449) for the HHL risk group. The mean (SD) values of the *left* AP amplitude in *women* were 0.91 (0.437) for the control group and 0.82 (0.399) for the HHL risk group. ANOVA showed that there was no significant difference among the four groups (F = 0.316, p = 0.814).

**FIGURE 4.**
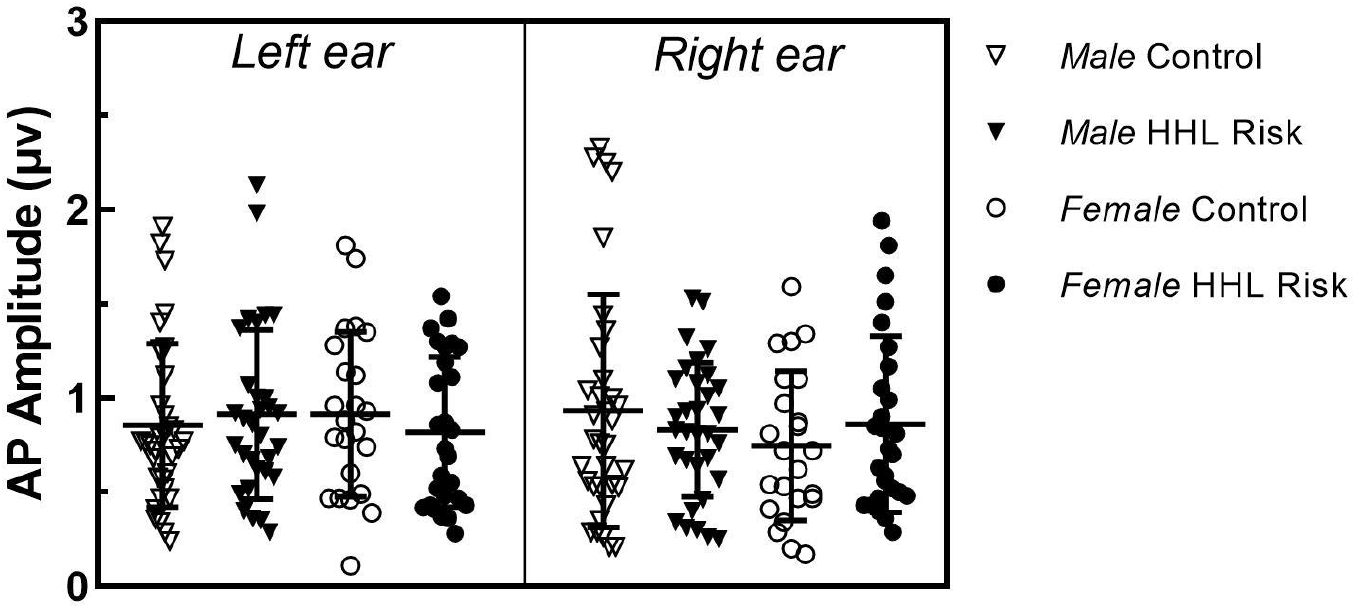
The AP amplitude of the control and the HHL risk group by sex. The AP amplitudes of both ears are shown in different panels. The AP amplitude further separated by sex was compared. No significant difference was found.

The mean (SD) values of the *right* AP amplitude in *men* were 0.94 (0.612) for the control group and 0.83 (0.355) for the HHL risk group. The mean (SD) values of the *right* AP amplitude in *women* were 0.75 (0.395) for the control group and 0.86 (0.468) for the HHL risk group. ANOVA showed that there was no significant difference among the four groups (F = 0.819, p = 0.486).

The AP latency values of both ears are plotted in Figure 5. The details of the *left* ear are plotted on the left, and those of the *right* ear are plotted on the right. The mean (SD) values of the *left* AP latency in *men* were 1.59 (0.124) for the control group and 1.59 (0.134) for the HHL risk group. The mean (SD) values of the *left* AP latency in *women* were 1.53 (0.113) for the control group and 1.57 (0.118) for the HHL risk group. ANOVA showed that there was no significant difference among the four groups (F = 1.444, p = 0.234).

**FIGURE 5.**
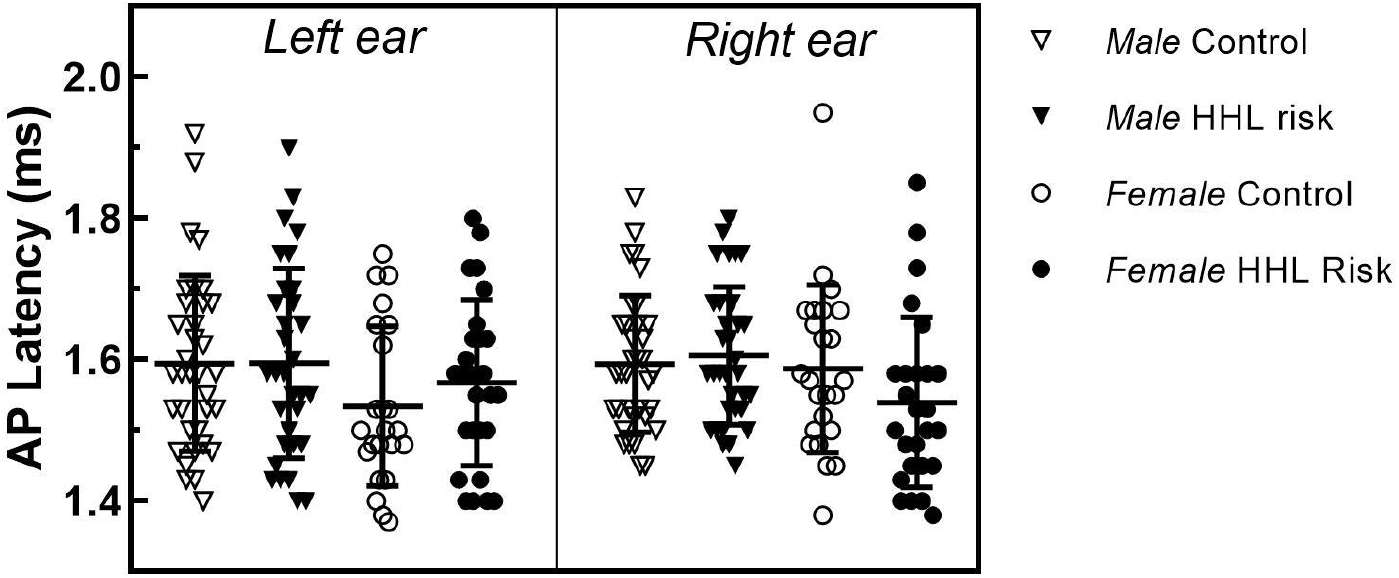
The AP latency of the control and the HHL risk group by sex. The AP latency of either ear is shown in different panels. AP latency further separated by sex was compared. No significant difference was found.

The mean (SD) values of the *right* AP latency in *men* were 1.59 (0.097) for the control group and 1.61 (0.098) for the HHL risk group. The mean (SD) values of the *right* AP latency in *women* were 1.59 (0.119) for the control group and 1.54 (0.120) for the HHL risk group. ANOVA showed that there was no significant difference among the four groups (F = 2.052, p = 0.111).

The SP/AP values of both ears are plotted in Figure 6. The details of the *left* ear are plotted on the left, and those of the *right* ear are plotted on the right. The mean (SD) values of the *left* SP/AP in *men* were 0.23 (0.062) for the control group and 0.30 (0.094) for the HHL risk group. The mean (SD) values of the *left* SP/AP in *women* were 0.25 (0.074) for the control group and 0.25 (0.062) for the HHL risk group. A nonparametric test showed that the difference among the four groups was significant (p = 0.006). A post hoc test showed a significant difference between the *men* (p = 0.004). The difference between the *women* was not significant (p = 1.000).

**FIGURE 6.**
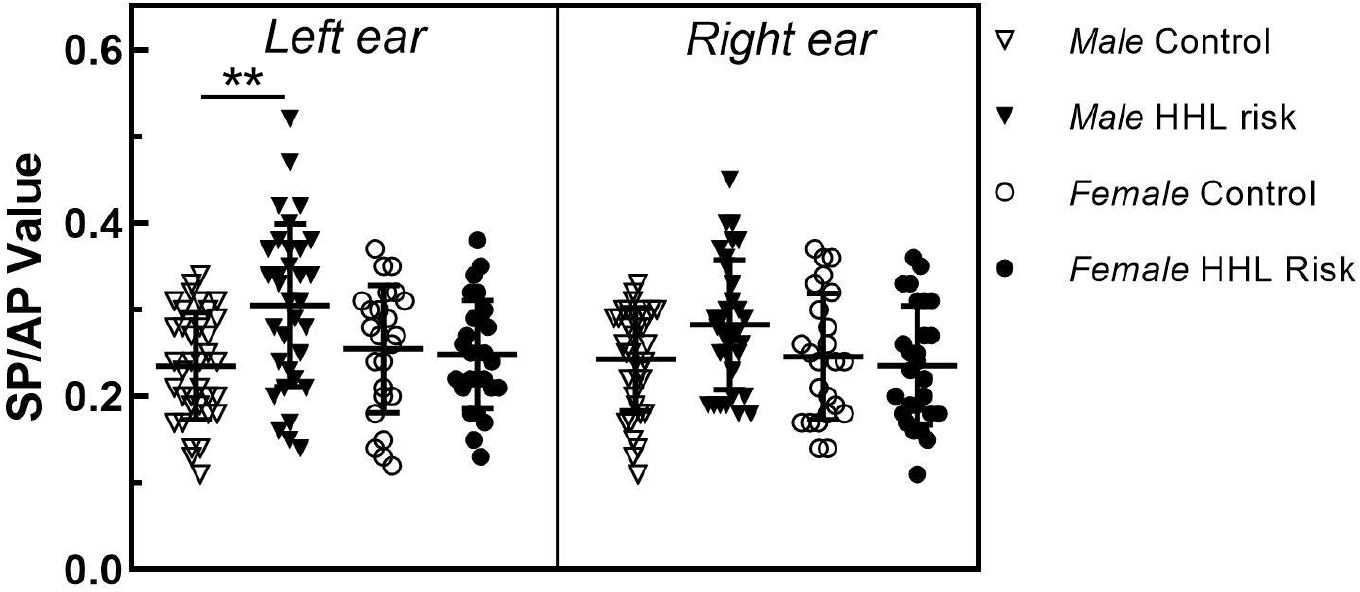
The SP/AP values in the control and HHL risk groups by sex. The SP/AP values of both ears are shown in different panels. SP/AP values further separated by sex were compared. Significance was only found in men for the left ear. The p values less than 0.01 are indicated **.

The mean (SD) values of the *right* SP/AP in *men* were 0.24 (0.059) for the control group and 0.28 (0.075) for the HHL risk group. The mean (SD) values of the *right* SP/AP in *women* were 0.25 (0.073) for the control group and 0.24 (0.068) for the HHL risk group. ANOVA showed that the difference among the four groups was significant (F = 2.912, p = 0.037). However, a post hoc test showed that the difference between neither the *men* (p = 0.112) nor the *women* (p = 1.000) groups was significant.

### Correlation analysis

#### Correlation of the SIN score and cochlear function

The correlation of the SIN score and cochlear function was explored by calculating Pearson correlations among 120 participants by sex. Significance was only found between the SIN score and the AP amplitude of the *right* ear in *men* (p = 0.034). The scatter plots of both sexes are displayed in Figure 7. No significant difference was found between the SIN score and cochlear function in *women*. Detailed results of the Pearson correlation analysis are shown in Table 1.

**FIGURE 7.**
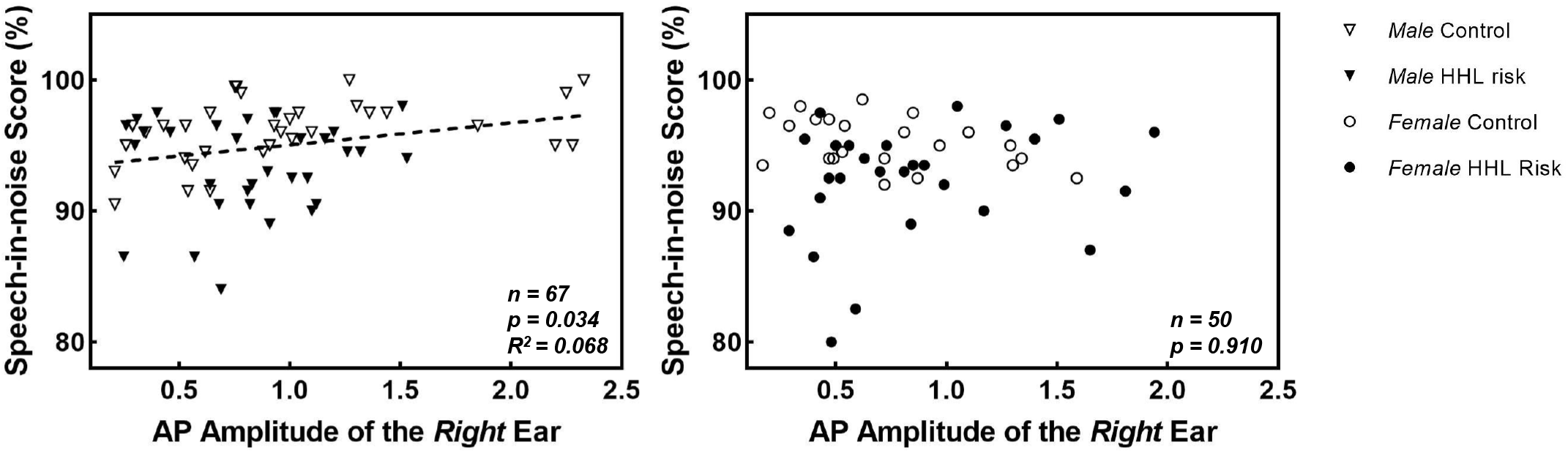
Scatter plot of the AP amplitude of the right ear and the SIN score. Male and female individuals are displayed separately in different panels. Regression lines are plotted for the men as dashed lines. The AP amplitude values of two male and one female participant were considered outliers and excluded.

### Relationship of OWL with auditory processing ability and cochlear function

The relationship of OWL with auditory processing ability and cochlear function was also explored by calculating Pearson correlations in occupational noise-exposed participants. However, a significant difference was only found between OWL and the cochlear function of the AP latency of the *right* ear (p = 0.002). Sex differences were further explored in the AP latency of the *right* ear. The results demonstrated that a significant correlation was only shown between OWL and the AP latency of the *right* ear in *men* (p = 0.025). The correlation between OWL and the AP latency of the *right ear* in *women* was not significant (p = 0.138). The scatter plots of both sexes are displayed in Figure 8. Detailed results of the Pearson correlation analysis are shown in Tables 2 and 3.

**FIGURE 8.**
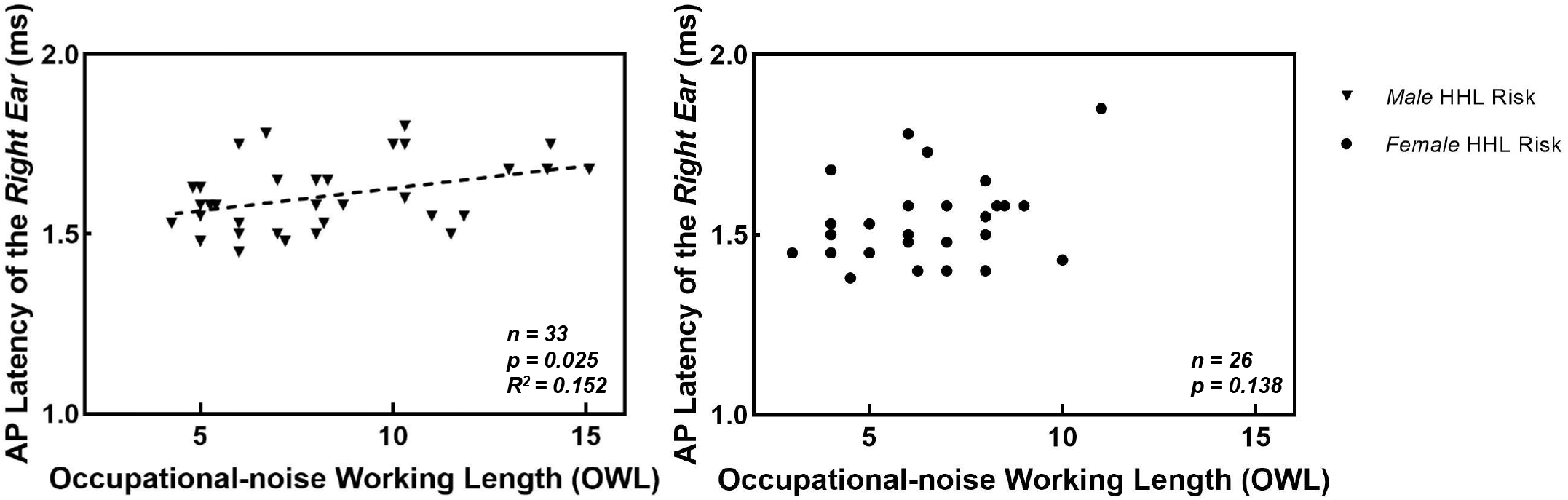
Scatter plot of OWL and the AP latency in the HHL risk group. Male and female individuals are displayed separately in different panels. Regression lines are plotted for the men as dashed lines. The AP latency value of one female participant was considered an outlier and excluded.

## Discussion

Our results demonstrated that although the hearing thresholds of the women in the HHL risk group were slightly but significantly worse than those of the control group, no significant differences in auditory processing ability, temporal resolution or cochlear function were shown. Sex differences in noise-induced hearing loss have been documented previously. One retrospective study of industrial factory workers reported that women experience less severe deterioration of hearing thresholds than men when occupational sound intensities are approximately 98 dB [19]. Female chinchillas display less permanent threshold shifts (PTS) than males at most frequencies after 150 dB peak sound pressure level noise exposure [20]. Studies have also shown that female mice are relatively protected from PTS-inducing noise exposure in comparison to males [21]. It has also been shown that men are at higher risk of high-frequency hearing loss than women despite equivalent noise exposure and age in another study in a population with occupational noise [22]. The women in our study displayed no significant disability of auditory processes, temporal resolution or cochlear function. The sex steroid milieu is believed to play an important role in sex differences in hearing physiology. Oestrogen levels are correlated with the age of women. In this research, the control and HHL risk groups were in the premenopausal period with similar ages.

Defects in the SIN scores and 4 kHz GDT were only significant in the HHL risk group in men. A classic symptom of auditory process disorder in humans with normal hearing is speech perception disability in a noisy background. Animal studies have suggested that CS could also play an important role in those populations. As a result, a variety of measures have been used to determine whether peripheral auditory dysfunction of noise-induced CS occurs in humans [11, 12, 16, 17, 23, 24]. However, postmortem temporal bone studies have demonstrated that CS and neural degeneration exist widely among humans [25, 26], even in young adults [27]. Studies have shown that other factors, including ageing [27-30] and ototoxic drugs [31, 32], also cause CS. In other words, CS is probably prevalent in young adults, so it is difficult to find significant differences between populations.

In our study, a higher SP/AP value was shown in men with long-term occupational noise exposure, but it was not correlated with the SIN score or OWL. This result suggested that CS differences could be found in populations with different noise exposure doses; however, the decreased ability of hearing in background noise in those populations was not related to CS. Hearing in background noise requires not only intact peripheral auditory function but also intact central auditory factors such as attention, working memory and language. CS could be one of the cochlear dysfunctions in humans. More recent findings indicated that other causes of HHL exist in humans, including cochlear demyelination [33, 34] and possibly mild or persistent hair cell dysfunction [35, 36]. Other studies have demonstrated that noise exposure also has a negative effect on the central auditory pathway. For example, Dewey et al. reported that fMRI responses throughout the auditory system were greater in individuals with higher lifetime noise exposure levels than in controls with low lifetime noise exposure levels [37]. In summary, the decreased SIN scores in men in the HHL risk group in our study could result from other dysfunctions of the cochlear or central auditory system.

A previous study showed that even among young people with normal audiograms, subclinical outer hair cell (OHC) dysfunction could affect the hearing ability in background noise [35, 38]. Previous studies reported lower levels of OAEs in association with hazardous noise-exposed populations even when hearing thresholds were within normal limits [39, 40] and levels of OAEs were significantly reduced in the second-year evaluation in a cohort with occupational noise exposure [41]. In our study, the OHC function of otoacoustic emissions (OAEs) was not measured. However, according to a previous study, individuals with occupational noise probably have significantly reduced levels of OAEs. However, the women in our study showed no significant decrease in SIN scores. This suggested that OHC dysfunction could appear earlier than CS in humans.

## Conclusion

In individuals with long-term occupational working exposure and normal hearing, defects in auditory processing, temporal resolution and cochlear function were observed only in men. In men, a weak correlation between the SIN score and the AP amplitude of the right ear was observed. There was only a weak correlation between OWL and the AP latency of the right ear. It appears that men are more vulnerable to occupational noise than women. Considering the noise-exposure dose differences between the control and HHL risk groups, our measures are insensitive to cochlear synaptopathy in humans.

## Data Availability

All data referred to in the manuscript are availability

## ACKNOWLEDGMENT

This research was supported by the National Natural Science Foundation of China (grant 81700903 to BL) and Shanghai Key Laboratory of Translational Medicine on Ear and Nose Diseases (14DZ2260300).

Bei Li, Zhiwu Huang and Hao Wu designed the study. Zhiwu Huang and Qixuan Wang recruited the participants. Qixuan Wang, Lu Yang and Yun Li collected the data. Bei Li and Qixuan Wang analyzed the data. Bei Li wrote the manuscript. All authors revised and edited the manuscript.

## List of abbreviations

HHL: Hidden Hearing Loss
SIN: Speech-in-noise
GDT: Gap Detection Threshold
AP: Action Potential
SP: Summating Potential
OWL: Occupational-noise Working Length
TTS: Temporal Threshold Shift
IHC: Inner Hair Cell
CS: Cochlear Synaptopathy
CAP: Compound Action Potential
ABR: Auditory Brainstem Response
FFR: Frequency-following Response
MHINT: Mandarin version of the Hearing in Noise Test
ANOVA: Analysis of Variance
PTS: Permanent Threshold Shifts
OHC: Outer Hair Cell
OAE: Otoacoustic Emissions

