## Supplementary material for "Characteristic and Sex Differences in Auditory Function and Cochlear Pathophysiology in a Noise-exposed Cohort: A Cross-sectional Study": Tables

**Table 1. Correlation analysis of the speech-in-noise score and cochlear function**

|  | *Left* Ear | | | *Right* Ear | | |
| --- | --- | --- | --- | --- | --- | --- |
|  | AP Amplitude | AP Latency | SP/AP Value | AP Amplitude | AP Latency | SP/AP Value |
| ***Male SIN score*** |  |  |  |  |  |  |
| *Pearson* correlation | -0.040 | -0.171 | -0.231 | 0.260* | -0.022 | -0.130 |
| P value | 0.751 | 0.166 | 0.056 | 0.034 | 0.861 | 0.290 |
| ***Female SIN score*** |  |  |  |  |  |  |
| *Pearson* correlation | 0.058 | -0.135 | -0.247 | 0.016 | 0.093 | -0.206 |
| P value | 0.691 | 0.350 | 0.081 | 0.910 | 0.521 | 0.147 |

The p values less than 0.05 are indicated*.

SIN is short for speech-in-noise

**Table 2. Correlation analysis of the occupational-noise working length with auditory processing ability**

|  | Speech-in-noise Score | 4 kHz Marker | 2 kHz Marker | 1 kHz Marker |
| --- | --- | --- | --- | --- |
| *Pearson* correlation | 0.173 | -0.119 | 0.041 | -0.067 |
| P value | 0.185 | 0.368 | 0.757 | 0.612 |

**Table 3. Correlation analysis of the occupational-noise working length with the cochlear function**

|  | *Left* ear | | | *Right* ear | | |
| --- | --- | --- | --- | --- | --- | --- |
|  | AP Amplitude | AP Latency | SP/AP Value | AP Amplitude | AP Latency | SP/AP Value |
| *Pearson* correlation | -0.019 | -0.053 | 0.196 | 0.077 | 0.399* | -0.077 |
| P value | 0.886 | 0.692 | 0.134 | 0.561 | 0.002 | 0.563 |

The p values less than 0.05 are indicated*.
